# Differences in prognostic value of *FLT3* and *NPM1* mutations in older and younger patient populations with acute myeloid leukemia

**DOI:** 10.1101/2025.11.16.25340236

**Authors:** Claudio A. Mosse, Michele LeNoue-Newton, Julie A. Lynch, Amy M. Perkins, Fern Fitzhenry, Kathryn M. Pridgen, Freneka Minter, Mia Levy, Michael E. Matheny

**Affiliations:** Department of Pathology, Microbiology, and Immunology, Vanderbilt University Medical Center, Nashville, TN, USA; Geriatric Research Education and Clinical Care Service & VETWISE-LHS COIN, Tennessee Valley Healthcare System (TVHS) VA Medical Center, Veterans Health Administration, Nashville, TN, USA; Vanderbilt Ingram Cancer Center, Vanderbilt University Medical Center, Nashville, TN, USA; VA Informatics and Computing Infrastructure, VA Salt Lake City Health Care System, Salt Lake City, UT, USA; Department of Epidemiology, University of Utah School of Medicine, Salt Lake City, UT, USA; Department of Biostatistics, Vanderbilt University Medical Center, Nashville, TN, USA; Foundation Medicine, Cambridge, MA, USA; Department of Biomedical Informatics, Vanderbilt University Medical Center, Nashville, TN, USA; Department of Medicine, Vanderbilt University Medical Center, Nashville, TN, USA

**Keywords:** *FLT3*, *NMP1*, acute myeloid leukemia, prognostic markers

## Abstract

The prognostic value of nucleophosmin-1 (*NPM1*) and fms-like tyrosine kinase 3 (*FLT3*) mutations has been well-established in adult patients but is less clear in geriatric patients. This retrospective cohort study assessed adult patients with acute myeloid leukemia (AML) who received *FLT3* and/or *NPM1* testing treated at any Veterans Health Administration (VHA) facility or at Vanderbilt University Medical Center (VUMC) between January 2006 and December 2016. The primary analysis compared time to all-cause death among patients with AML based on *FLT3* and *NPM1* mutation status, age (<65 years, ≥65 years), and cytogenetic risk group (unfavorable, intermediate/normal, favorable). The study population (n=766) (mean age 59.71 ± 16.6 years, 45.96% ≥65 years) had a mutation rate of 19.8% for *FLT3* and 22.1% for *NPM1*. Age was a significant factor in overall survival (median OS: 24.28 vs. 8.18 months, p<0.001), as was cytogenetic risk status (median OS: favorable group not reached, intermediate/normal 17.61m vs. unfavorable 6.77m, p<0.001). The most favorable prognostic group (*FLT3*-/*NPM1*+) among older patients showed worse OS (15.21 months) than did the poor prognostic group (*NPM1*-/*FLT3*-) among younger patients (18.43 months). Among the older cohort, *FLT3* and *NPM1* mutation status, favorable karyotype, and CCI were not identified as prognostic factors. In the full cohort, using Cox proportional hazard regression and LASSO analyses for age, *FLT3* and *NPM1* mutation status, cytogenetic risk group, treatment site, race, primary payor, and Charleston Comorbidity Index (CCI), age (HR 65y vs 35y/=2.44, 95% CI 1.61-3.68, p<0.001) was the strongest risk factor in AML.

## Introduction

Acute myeloid leukemia (AML) is an aggressive hematologic malignancy that, when left untreated, rapidly leads to death. Over 20,000 new cases of AML are diagnosed every year within the US,^1^ with incidence rates of AML increasing with age from less than 2 per 100,000 in patients under 65 to over 16 per 100,000 in patients over 65 years of age.^2^ Over 57% of newly diagnosed AML occurs in patients over 65 with over one-third of new cases in patients over 75 years old.^3^ In addition to higher incidence rates, patients over 65 also have worse outcomes and lower 5-year survival rates, 7% compared to approximately 45% in those under 65.^3^ While age itself is an important independent prognostic risk factor for survival and response to treatment,^4–7^ the low 5-year survival rates for patients over 65 is influenced by numerous other factors including decreased performance status,^8^ increased rates of unfavorable cytogenetic and molecular risk factors,^4,9^ as well as decreased usage of intensive chemotherapy and bone marrow transplant. ^10–12^

Cytogenetic and molecular risk factors are important prognostic factors for survival and factor into decisions on post-remission therapy.^13^ Cytogenetics is used to stratify patients into high, intermediate, and low risk populations, and unfavorable cytogenetics is a predictor of poor outcomes across all age groups.^9,13–15^ Normal karyotype is classified within the intermediate risk category and accounts for approximately 40% of the population; the heterogeneous nature of this group has led to further stratification based upon molecular abnormalities in FMS-like tyrosine kinase (*FLT3*) and nucleophosmin-1 (*NPM1*).^10^

*FLT3*-ITD and *NPM1* are two common molecular biomarkers occurring in approximately 21% and 36% of all patients with AML, respectively.^16,17^ *NPM1* and *FLT3* (ITD and TKD) mutations are all more frequent in patients with normal karyotype and de novo AML,^18–23^ with *FLT3* mutations twice as common in patients with the *NPM1* mutation than in patients with *NPM1* wildtype.^18,24^ *NPM1* mutations are considered a favorable prognostic factor associated with increased rates of complete remission (CR) and superior overall survival (OS) and disease-free survival (DFS), at least in the absence of co-occurring *FLT3*-ITD mutations.^25–28^ In contrast, *FLT3*-ITD mutations are considered a poor prognostic factor with decreased OS and DFS, though co-occurrence with *NPM1* mutations improves outcomes over *FLT3*-ITD alone.^16,22,23,29^ The effects of *FLT3*-TKD mutations have been mixed with studies showing decreased OS and DFS with TKD mutation alone or in combination with ITD,^16,22,30,31^ but paradoxically improved OS when in combination with *NPM1* mutations compared to the presence of the *NPM1* mutation alone.^24,29,30^

While well-established in the younger patient population, the prognostic significance of *NPM1* and *FLT3*-ITD within geriatric patients is less clear, although recent studies have examined this within various AML study groups and registries.^32,33^ This current study uses a large real-world cohort of adult patients with AML treated at either Veterans Health Administration or an academic institution to investigate the prognostic implications of *NPM1* and *FLT3*-ITD mutations in those over 65 years old compared to patients under 65 years old.

## Methods

### Study Population

The study population consisted of patients diagnosed with AML between January 1, 2006 and December 31, 2016, who sought care at Vanderbilt University Medical Center (VUMC) or within any Veterans Health Administration (VHA) facility nationally. Patients met the following inclusion criteria: 1) age 18 years or older, 2) newly diagnosed or initially treated at the respective institution and documented in the cancer registry and 3) received *FLT3* and/or *NPM1* testing.

### Identification of Tests and Test Results

Within this cohort of patients, molecular tests for *FLT3*-ITD and *NPM1* mutations were identified along with dates of testing. Cytogenetics were classified as Favorable, Unfavorable, Intermediate or Normal, as per NCCN Risk Category guidelines.

At VUMC, the relevant results and testing dates from the Enterprise Data Warehouse (EDW) were extracted. Next-generation sequencing (NGS) tests were also identified and results for *FLT3*-ITD, *FLT3*-TKD, and *NPM1* mutations were manually abstracted from the results. We conducted further review of patient’s electronic health record (EHR) for those patients without testing information at diagnosis to identify any outside testing or unstructured test results.

At the VHA, structured test results and testing information were taken from the national GDX database. We identified additional patients with *FLT3*-ITD or *NPM1* testing by test name and keyword searches of notes and test results within the EHR. Test results for *FLT3*-ITD, *FLT3*-TKD, *NPM1* genomic mutations and cytogenetic results were abstracted either from test comments or from manual review of EHR.

### Determination of Diagnostic Testing

We defined diagnostic molecular and cytogenetic tests as test results received within 14 days prior to the date of diagnosis or within 12 days post-date of diagnosis. Any patient without test results within this range and some indication that testing was conducted on a sample taken within the range underwent manual chart review to identify possible diagnostic test results (e.g., molecular testing that resulted 4 weeks post-date of diagnosis, but lab report states the sample was a diagnostic sample). Test results falling outside of this range were reviewed and included as diagnostic test results if one of the following conditions was met: 1) test was performed no more than 30 days prior to date of diagnosis, 2) test was performed within 30 days of diagnosis with residual disease, defined as >20% blast for Normal/Not Detected results, 3) test was performed within 60 days of diagnosis in untreated disease, or 4) test was performed within 60 days of initial diagnosis and within 30 days of completion of initial treatment with residual disease, defined as >20% blasts for Normal/Not Detected.

### Statistical Analysis

The primary analysis compared time to all-cause death among patients with AML who tested positive or negative for either *FLT3* mutations or *NPM1* mutations. Time 0 was the date of AML diagnosis. Patients were censored at last patient contact or end of the study period (12/31/2016). Seventy-eight patients who did not have both *FLT3* and *NPM1* genetic tests were excluded from analyses. Missing data were imputed using multivariate imputation by chained equations (MICE). Kaplan-Meier plots and logrank tests were reported by age group (<65 years, ≥65 years), *FLT3* and *NPM1* status (+/-), and cytogenetic group (unfavorable, intermediate/normal, favorable). We performed a penalized Cox proportional hazards (PH) regression, using the L1 penalty (least absolute shrinkage and selection operator [Lasso]) to select a subset of predictor variables. The tuning parameter λ, which controls the strength of the penalty, was selected based on cross-validation (CV) using the mean square error as the criterion. We selected the λ with the minimum CV error. We subsequently used the variables identified by the Lasso procedure (e.g., age at diagnosis, race, cytogenetic group, primary payor, and Charlson Comorbidity Index^34^ [CCI]) and variables identified a priori *FLT3* status, *NPM1* status, and site) in an unpenalized Cox PH model. Restricted cubic splines with 3-5 knots were used for all continuous variables. Linear contrasts for age and CCI were also reported. Preplanned subgroup analyses were performed by stratifying based on age (<65 vs. ≥65). All statistical analyses were conducted using R software version 3.5.1.^35^

## Results

### Patient Demographics

A total of 439 patients with AML from VUMC and 327 from the VHA received testing for either *FLT3* or *NPM1*, resulting in a total study population of 766 patients. The overall population had a mean age at diagnosis of 59.71 ± 16.62 years and was predominantly white (84.5%) and male (74.7%). Insurance coverage of the patients was mostly either private insurance (25.5%), Medicare (31.6%) or military/Tricare (32.4%). Most VHA patients were diagnosed at VHA facilities rated either 1a (62.1%) or 1b (22.6%) on the VHA hospital complexity scale where the options, from most to least complex, are 1a, 1b, 1c, 2, and 3. The mean distance between the patient and diagnostic medical center was 114.70 ± 186.32 miles, and the average distance from the diagnostic medical center to an NCI designated cancer center was 26.62 ± 67.62 miles. Most of the patients had prior diagnoses of either myelodysplastic syndrome (MDS) (29.2%) or myeloproliferative neoplasm (MPN) (26.2%).

### Site Differences

There was a significant difference in mean age at diagnosis between the VUMC and VHA patient populations (VUMC: 55.84 years, VHA: 64.90 years; p<0.001) and in gender distribution (VUMC: 57.9% male, VHA: 97.2% male; p<0.001). The VUMC population had a slightly lower percentage of non-White patients (12.5% vs. 19.6%) (p=0.01) and had lower co-morbidities as measured by CCI (VUMC: 0.40, VHA: 1.43; p<0.001). The VUMC patient population resides in areas with lower rates of college-educated individuals (21.30% VUMC vs. 27.80% VHA, p<0.001) and higher rates of persons below the poverty level threshold than the VHA patient population (17.53% VUMC vs. 15.76% VHA, p<0.001). The VUMC patient population was much more likely to have a reported prior hematological neoplasm with MDS (VUMC: 42.8%, VHA: 11.0%; p<0.001) and MPN (VUMC: 42.6%; VHA: 4.3%; p<0.001) being the most prevalent prior conditions noted.

### Age-Stratified Cohort Differences

The population was roughly evenly distributed between 414 patients younger than 65 years old and 352 patients 65 years old or older. The average age of the <65 years cohort was 49.25 ± 15.62 years, and the average age of the ≥65 years cohort was 72.01 ± 5.81 years. We observed differences in gender distribution between the age-stratified cohorts (<65 years: 68.4% male; ≥65 years: 82.1% male). There were no significant differences in racial distribution between patients aged <65 years and patients >=65 years old. Patients aged >=65 years old had greater co-morbidities as measured by the CCI (<65 years: 0.60, ≥65 years: 1.12; p<0.001). The older cohort resided in areas with higher rates of college-educated individuals (<65 years: 23.09% vs. ≥65 years: 25.23%; p=0.004) and lower rates of persons below the poverty level threshold (<65 years: 17.05%, ≥65 years: 16.45%; p=0.04) than the younger cohort. The older cohort was much more likely to have previously reported MDS (<65 years: 25.1%, ≥65 years: 34.1%; p=0.01) and less likely to have previously reported MPN (<65 years: 31.9%, ≥65 years 19.6%; p<0.001).

### Mutation Frequency and Cytogenetics

The combined population had a 19.8% rate of *FLT3* mutation and a 22.1% rate of *NPM1* mutation, with 59.3% of patients being double negative (*FLT3*-/*NPM1*-) and 10.2% being double positive (*FLT3*+/*NPM1*+). The younger cohort was more likely to be *FLT3*-positive (<65 years: 24.9%, ≥65 years: 13.9%; p<0.001) with no differences detected between age cohorts in rates of *NPM1* positivity. Younger patients were also more likely to be double positive than older patients (<65 years: 13.3%, ≥65 years: 6.5%; p=0.002) and less likely to be solely *NPM1*+ (<65 years: 9.2%, ≥65 years: 14.8%; p=0.02). Of note, VUMC patients were more likely to have *FLT3* mutations than VHA patients (VUMC: 25.3%, VHA: 12.5%; p<0.001). Older patients were more likely to have unfavorable cytogenetics (<65 years: 26.3%, ≥65 years: 35.2%; p=0.01) and less likely to have favorable cytogenetics (<65 years: 9.2%; ≥65 years: 2.3%; p<0.001).

### Overall Survival Analysis

In an unweighted, unadjusted analysis, age was a significant factor in overall survival (OS) amongst patients with AML, with older patients showing significantly worse outcomes (median OS: 24.28 months vs. 8.18 months, p<0.001) (Figure 1). Cytogenetic risk status was also a major factor in overall survival amongst patients with AML (median OS: not reached in the favorable group, 17.61 months for intermediate/normal, 6.77 months for unfavorable, p<0.001), an influence that held within the age-stratified cohorts (Figure 2). Although cytogenetics did have an influence on overall survival in the older cohort, the effect was diminished (median OS: 9.69 months for the favorable group, 9.82 months for the intermediate/normal, 5.22 months for the unfavorable, p=0.001). In fact, patients in the older cohort with favorable cytogenetics showed roughly equivalent survival to younger cohort patients with unfavorable cytogenetics (9.69 months vs 8.25 months).

**Figure 1:**
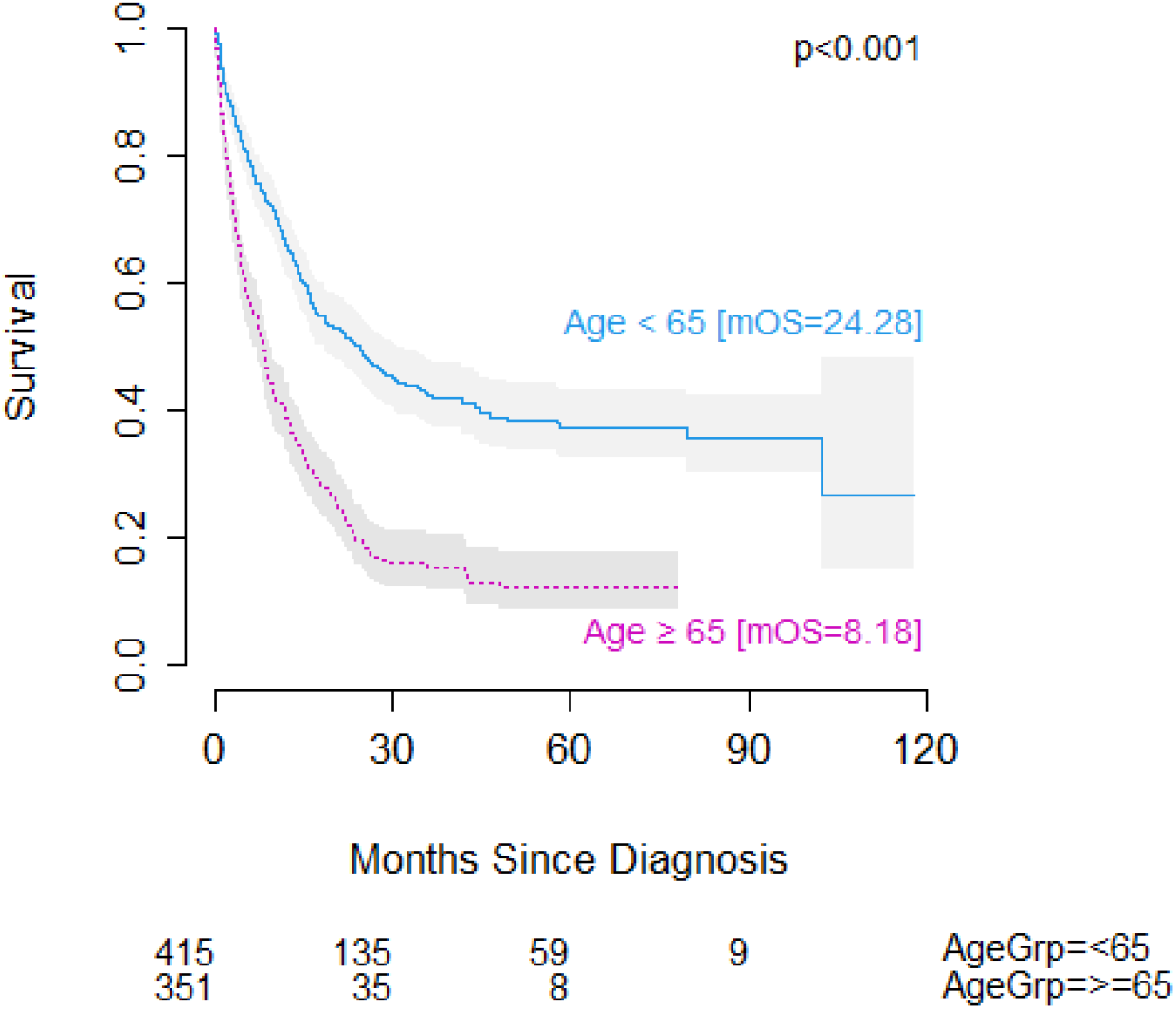
Overall survival in patients with AML <65 years of age vs. ≥65 years of age. Number at risk shown below. mOS = median overall survival.

**Figure 2.**
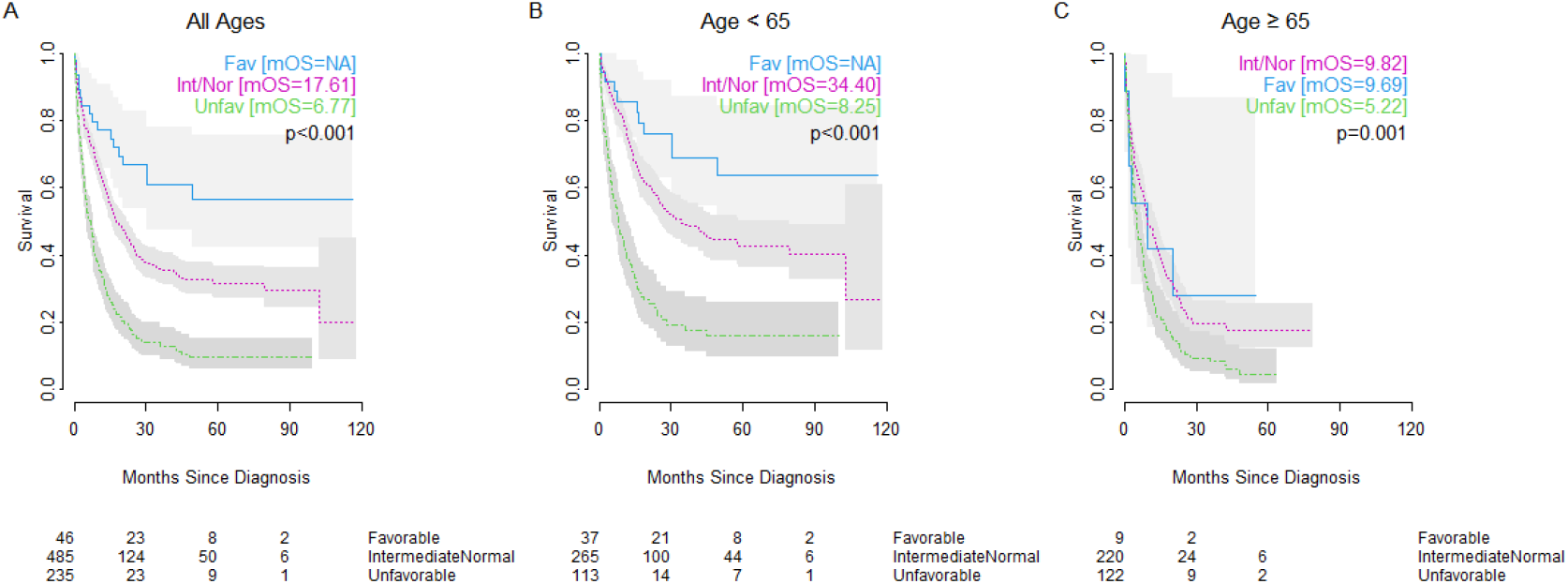
Impact of cytogenetics on overall survival in patients with AML of (A) all ages, (B) age <65 years, or (C) age ≥65 years of age with favorable cytogenetics (blue solid line), intermediate/normal cytogenetics (pink dotted line), or unfavorable cytogenetics (green dashed line). Number at risk shown below. mOS = median overall survival.

*NPM1* and *FLT3* mutations were prognostic factors for OS when looking at co-mutation groups. Across all ages, we observed that those with *NPM1* mutation in the setting of wild-type *FLT3* had the most favorable outcomes, with significantly better outcomes than even patients with wild-type *NPM1* and wild-type *FLT3* (double-negative) (Figure 3A) (25.79 months vs 12.71 months). However, this effect was driven almost entirely by younger cohort (<65 years) (Figure 3B) with little difference in OS seen between mutation groups in the older cohort (≥65 years) (Figure 3C). Although not reaching significance, we observed a similar pattern in the subset of patients with normal/intermediate risk cytogenetics, where the trend of differences in overall outcomes was also driven by the younger cohort. (Figure 4).

**Figure 3.**
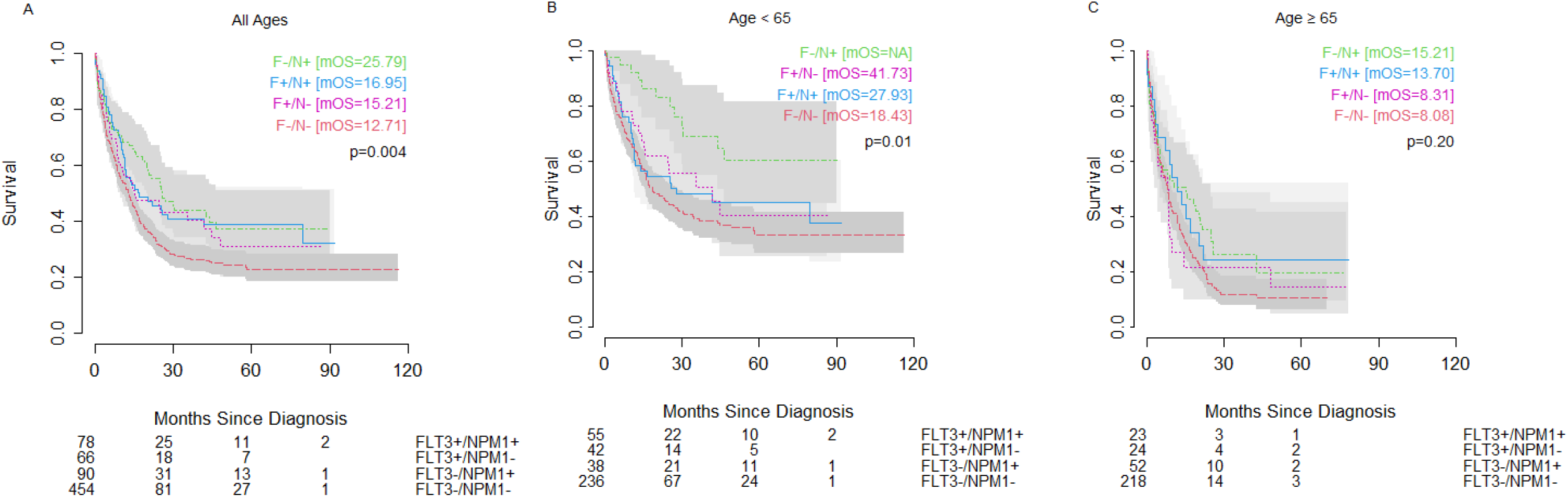
Impact of *FLT3* and *NPM1* on overall survival in patients with AML of (A) all ages, (B) age <65 years, or (C) age ≥65 years of age with wild-type *FLT3* and *NPM1* mutation (green dashed line), wild-type *FLT3* and wild-type *NPM1* (red dashed line), *FLT3* ITD or TKD and wild-type *NPM1* (pink dotted line), or *FLT3* ITD or TKD and mutated *NPM1* (blue solid line). Number at risk shown below. mOS = median overall survival.

**Figure 4.**
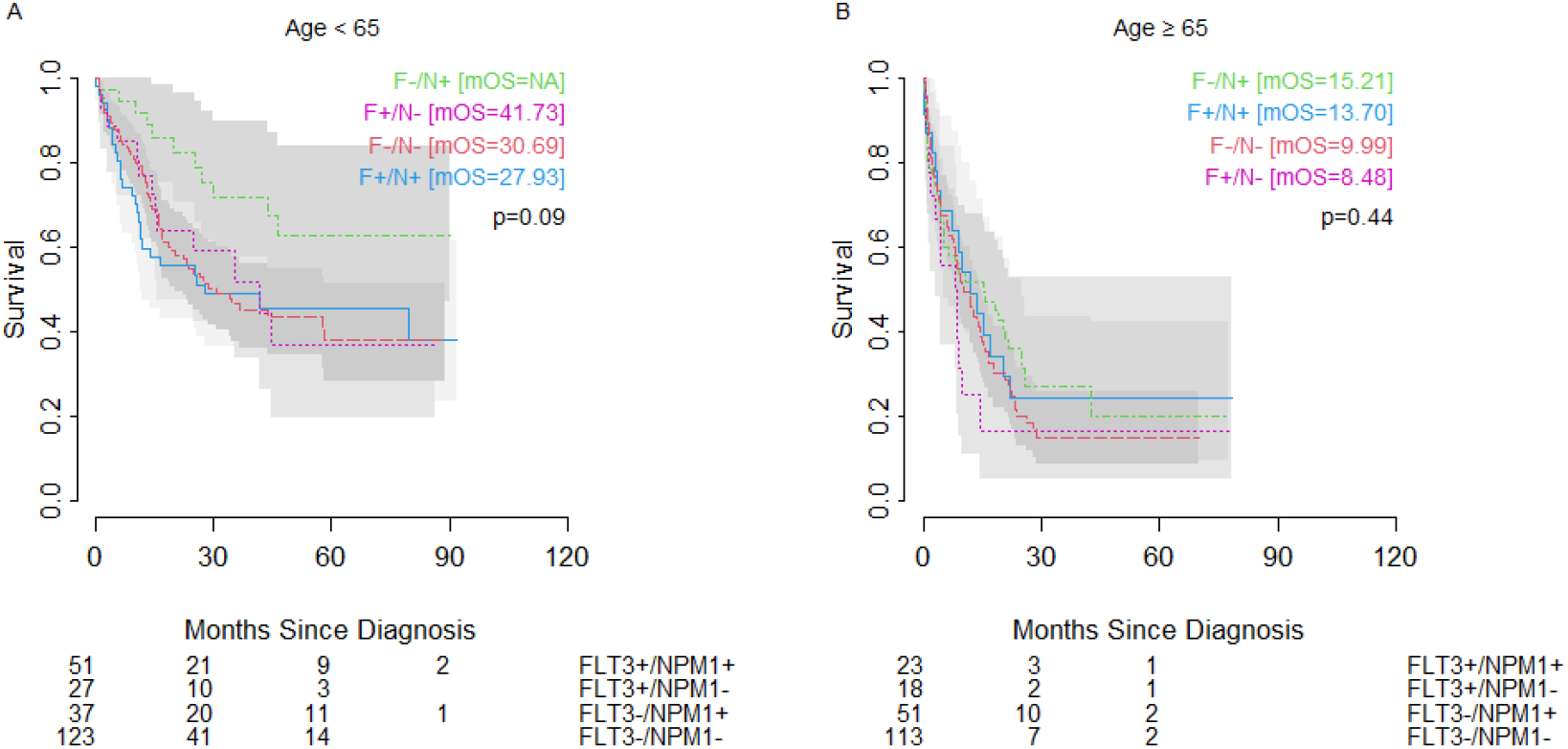
Overall survival in patients with intermediate/normal karyotype AML and (A) age <65 years, or (B) age ≥65 years of age with wild-type *FLT3* and *NPM1* mutation (green dashed line), wild-type *FLT3* and wild-type *NPM1* (red dashed line), *FLT3* ITD or TKD and wild-type *NPM1* (pink dotted line), or *FLT3* ITD or TKD and mutated *NPM1* (blue solid line). Number at risk shown below. mOS = median overall survival.

Of note, in both whole group analysis and when restricted to normal/intermediate karyotype, the most favorable prognostic group (*FLT3*-/*NPM1*+) among older patients showed a worse OS (median OS=15.21 months) than did the poor prognostic group (*NPM1*-/*FLT3*-) among younger patients (median OS=18.43 months). In both older and younger patients with unfavorable cytogenetics, patients with *FLT3*+ status showed better OS than patients with wild-type *FLT3*, though the difference was only significant within the younger patient population (Figure 5). The unfavorable cytogenetic risk group had no *NPM1*+/*FLT3*-patients in the <65 year age cohort and no *NPM1*+/*FLT3*+ patients in the ≥65 year age cohort.

**Figure 5.**
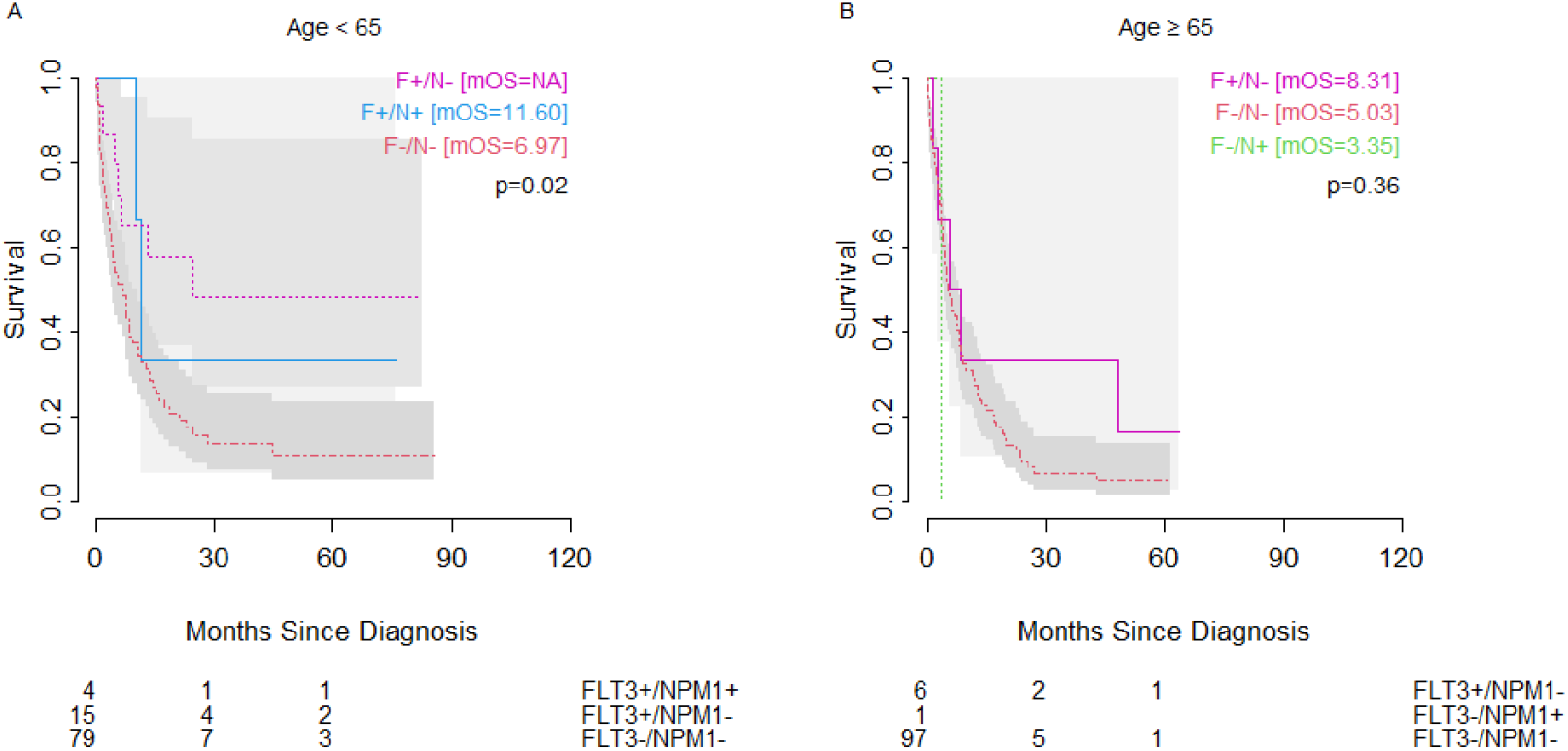
Overall survival in patients with unfavorable karyotype AML and (A) age <65 years, or (B) age ≥65 years of age with wild-type *FLT3* and *NPM1* mutation (green dashed line), wild-type *FLT3* and wild-type *NPM1* (red dashed line), *FLT3* ITD or TKD and wild-type *NPM1* (pink dotted line), or *FLT3* ITD or TKD and mutated *NPM1* (blue solid line). Number at risk shown below. mOS = median overall survival.

### Multivariable Cox Proportional Hazards Analysis

In multivariable Cox proportional hazards analysis, age at diagnosis, unfavorable cytogenetic risk status, higher CCI scores, and prior MPN diagnosis were selected by the LASSO method as having a significant effect on outcomes in the combined cohort of all ages. Age at diagnosis showed negative prognostic value (HR 65y vs 35y=2.44, 95% CI 1.61-3.68; p<0.001), as did unfavorable cytogenetics (HR=1.68, 95% CI 1.34-2.11; p=0.001) and CCI score (HR 2 vs 0=1.51, 95% CI 1.16-1.98; p=0.003). Prior MPN diagnosis was a favorable prognostic feature (HR=0.51, 95% CI 0.40-0.67; p<0.001). While not reaching significance in the multivariable analysis, *FLT3* mutation trended towards a negative prognostic value (HR=1.25, 95% CI 0.96-1.62; p=0.10) and *NPM1* mutation trended towards a positive prognostic value (HR=0.81, 95% CI 0.62-1.05; p=0.11).

In a multivariable analysis restricted to the younger cohort of patients (<65 years), unfavorable cytogenetic risk status and CCI score were negative prognostic factors and prior MPN diagnosis had positive prognostic significance. In this younger population, LASSO also identified treatment site at the VHA as positive prognostic factor (HR=0.48, 95% CI 0.26-0.89; p=0.02). In the older cohort, prior MPN diagnosis was again a favorable prognostic feature (HR=0.53, 95% CI 0.37-0.77; p<0.001) and Medicaid as the primary payor was a negative prognostic feature (HR=3.67, 95% CI 1.32-10.17; p=0.02). Neither *NPM1* mutation status (p=0.48), favorable karyotype (p=0.54) nor CCI (p=0.13) were selected as prognostic factors for OS in the older cohort. Also, in the older cohort *FLT3* mutation trended to a worse prognosis (HR=1.39, 95% CI 0.94-2.06) but did not reach statistical significance (p=0.10). Within an age-stratified cohort, age was a significant factor in overall survival, and older age was associated with increased hazard for death (HR 85y vs 65y=2.73, 95% CI 1.46-5.11; p=0.002).

## Discussion

Age, cytogenetics, and antecedent hematologic malignancy or previous history of chemotherapy have long been recognized as prognostic factors in AML. Advanced age has also been identified as a poor prognostic factor, either reflecting increased co-morbidities in the elderly or due to a distinct biology in elderly AML.^36,4,8,37,38^ Various recurrent molecular mutations have been identified as important prognostic factors as well.^39^ In particular, *FLT3*-ITD mutations are considered poor prognostic factors, particularly in the setting of wild-type *NPM1*. Many of the data used to determine the potential impact of *FLT3* and *NPM1* mutations have been derived from younger patients who are better able to withstand intensive induction chemotherapy than elderly patients are. Our current study examined the age-dependent prognostic factors on overall survival from an unrestricted nationwide cohort of 327 veterans treated at the VHA and 439 patients treated at an academic medical center. This study of US patients diagnosed and treated in both an academic medical center and in the national Veterans Health Administration represents one of the largest examinations of molecular prognostic factors in the elderly population of patients with AML.

This study examined the prevalence and prognostic impact of *NPM1* and *FLT3* mutations in a population of patients with AML. The older patients were statistically more likely to have a higher CCI, antecedent MDS, and unfavorable karyotype. The younger patients were more likely to have had an antecedent MPN. The older cohort showed a similar rate of *NPM1* mutations as the younger cohort. The prevalence of *NPM1* and *FLT3* mutations in this study is significantly different than that typically reported from younger AML cohorts^40,41^ but mirrors the findings seen in other studies of elderly patients with AML.^42,43^

In general, the older AML cohort has a decreased prevalence of *FLT3* mutations.^44^ While *FLT3*-ITD is generally considered a poor prognostic factor, the older cohort showed worse OS, both overall and in each of the four mutation subgroups. This was not merely a reflection of the increased rates of unfavorable karyotype among the elderly as the lack of prognostic impact of *FLT3* and *NPM1* was also observed when excluding patients with an unfavorable karyotype. In fact, when considering outcomes among patients ≥65 years old, favorable karyotype, *NPM1* status, nor *FLT3* status were statistically significant prognostic factors in a multivariable analysis. Among older patients with AML, Medicaid as a payor was a poor prognostic factor, likely reflecting impact of lower socioeconomic status on outcome. Interestingly, in the younger and older cohorts, antecedent MPN was a positive prognostic factor, possibly reflecting the widespread and highly effective use of BCR-ABL inhibitors in patients with AML with BCR-ABL translocations or perhaps a generally slower course of AML in diseases with antecedent MPN. In younger patients, comorbidities and favorable karyotype were still poor prognostic factors, as expected.^13^ Moreover, in the younger cohort, therapy within the VHA was a favorable prognostic factor, possibly reflecting the fact that VUMC acts as a regional referral center attracting therapy-resistant patients that would normally be treated in the community. Again, in the younger cohort, *FLT3* and *NPM1* mutations were not significantly associated with outcome when considered individually, although *NPM1*+/*FLT3*-patients trended to the best outcome and *FLT3*-/*NPM1*-patients trended towards a worse outcome in younger patients.

In this study, we observed that among patients with AML 65 years old or older, who comprise the majority of all patients with AML, the prognostic impact of favorable karyotype, *FLT3* mutations and *NPM1* mutations on OS is not statistically significant. Although numerous studies have shown a prognostic impact of *FLT3* and *NPM1* mutation status,^5,18,24,40,45^ those studies typically underrepresented older patients or excluded patients who could not undergo intensive induction chemotherapy. Other studies focusing on older patients with AML show similar findings to our current study, although often with smaller cohorts.

Scholl et al examined 99 elderly patients with AML (≥60 years), of whom 23 had *NPM1* mutations and 16 had the *FLT3*-ITD mutation.^46^ While *NPM1* status had no impact on OS, even when restricted to the *FLT3*-ITD-negative subset, patients with *FLT3*-ITD mutations treated with curative therapy (7 patients) showed a shorter OS (634d vs. 210d, p=0.03) than those without *FLT3*-ITD mutations (47 patients). In another study, Daver et al. examined elderly patients with AML (≥65 years old) with *NPM1* (146 patients) and *FLT3* testing (388 patients).^43^ While neither mutation alone showed statistically significant impact on OS in the elderly AML cohort, the *NPM1*-mutated/*FLT3*-wild type subset (14 patients) showed extended OS (21.5 months) compared to all others (9.0 months). Finally, a review of 156 patients treated on SWOG protocols failed to find a significant prognostic impact from *NPM1* or *FLT3* mutations, either separately or in combination, in the elderly AML cohort (≥65 years old).^42^ These same authors also reviewed outcomes from a cohort of 1258 UK Medical Research Council/National Cancer Research Institute (MRC/NCRI) patients, which included 448 patients older than 65 years and 810 patients between 55 and 65 years old. *NPM1*-mutated/*FLT3*-wild type patients older than 65 years showed no statistically significant improved OS compared to all other molecularly define subsets, whereas the younger patients with *NPM1*-mutated/*FLT3*-wild type AML did show improved OS as predicted from earlier studies on younger patients with AML. Juliusson et al^33^ studied 1570 adult patients on the Swedish AML registry and identified differences in the prognostic value of *NPM1* and *FLT3* among older (60-74 years) and younger (<60 years) patients. *FLT3*-ITD was a marker for poor prognosis in younger patients (p=.00003) but not in older patients. However, in contrast to our findings, *NPM1* mutation was a positive prognostic marker only in the older population (p=.00002). An age-stratified analysis by Straube et al^32^ observed that a *NPM1*+/*FLT3*-ITD-low status confers a poorer prognosis than *NPM1*+/*FLT3*-status younger patients (<60 years) but not in older patients (≥60 years), in which both mutation groups have similarly poor outcomes.

This current study affirms these previous real-world analyses that show little or no impact of *FLT3* and *NPM1* mutations among elderly patients with AML. Moreover, this study extends those previous findings by including karyotype classification to show that *FLT3* and *NPM1* status show no impact even within karyotype subclasses in elderly patients with AML. Notwithstanding our current findings and those from previous studies, the NCCN and ELN guidelines still define AML risk groups based on molecular findings without a recognition that patients over 65 are likely to do poorly regardless of mutation status and irrespective of co-morbidities. Moreover, as elderly individuals comprise the majority of patients with AML, the guidelines may better serve the patient population if they were structured primarily on age and then considered special circumstances in younger patients with different mutations.

There are several limitations to any observational study. First, over a 10-year time horizon, therapy was variable between patients, and likely influenced by patient presentation, age, and co-morbidities. We believe that younger patients were more likely to receive intensive induction therapy, and older patients were likely to receive less intense regimens. Unfortunately, we do not have therapeutic data available to confirm that hypothesis. Also, this data cannot account for changes in therapeutic paradigms over time. Increased use of demethylating agents in patients with MDS and in the elderly population of patients with AML were not assessed in this study. The use of midostaurin or other kinase inhibitors may have varied throughout the study although the time period analyzed predates widespread use of *FLT3*-specific inhibitors. Additionally, our data do not reflect the newer treatment regimens that have improved outcomes in elderly patients with AML. Moreover, the survival data was not censored at stem cell transplant for the minority of patients who may have received one. While stem cell transplant is unlikely to have been prevalent among the elderly cohort, among the younger patients, stem cell transplantation in the poor prognostic subgroups may have led to improved outcomes giving those younger, high-risk patients outcomes similar to intermediate risk patients and obscuring the negative impact of poor prognostic factors such as *FLT3* mutation and unfavorable cytogenetics.

In addition, there may be some limitations in the predominantly male and older VHA cohort, although patient features were adjusted when measured. As the VHA patients tended to skew older, and as antecedent MDS has been shown to be a poor prognostic factor, underreporting MDS in the VHA cohort would worsen the predicted outcome of the older cohort, which may explain why the older patients did so much worse across all molecularly defined subgroups. However, if there were significant underreporting of MDS among VHA patients, receiving therapy at the VHA, particularly for older patients, should have been a poor prognostic factor. In fact, older VHA patients did not have a worse outcome than those treated at the academic center, and in the younger cohort, treatment at the VHA was a good prognostic factor (HR=0.48, 95% CI 0.26-0.89; p=0.02). The worse outcome of younger patients at the academic center may be reflective of its role as a referral center for the most challenging patients for non-academic, community hematologists.

In conclusion, these results suggest that the patient’s age is the most significant prognostic factor in AML. Our findings may be complicated by the variability of therapy used in the elderly, but that variation in treatment reflects the biologic reality that these patients are often not able to tolerate the same treatments as younger patients. In essence, by not restricting this analysis to the healthiest of the elderly or to a population that skews significantly younger than the typical AML patient, our results reflect the grim reality facing physicians who care for older patients with AML– that there are no good prognostic groups. In an era burgeoning with new therapies, we must be cognizant of the complete spectrum of leukemia patients rather than developing prognostic algorithms and focusing trials primarily on the youngest and healthiest patients.

## Data Availability

VHA data is available to all VA researchers with appropriate IRB approvals.

## Acknowledgements

This work was supported through funding, access and computational resources, or facilities access by the following sources: 5-U01-HG-010232 Integrated, Individualized, and Intelligent Prescribing (I3P) Clinical Trial Network, Department of Veterans Affairs (VA) Informatics and Computing Infrastructure (VINCI), which is funded under the research priority to Put VA Data to Work for Veterans (VA ORD 24-D4V-02), and Veterans’ Wellbeing through Innovation, Systems Science and Experience in Learning Health Systems (VETWISE-LHS) (CIN-24-128). This publication does not represent the views of Vanderbilt University Medical Center or the Department of Veterans Affairs or the United States Government.

